# Surgical Pleth Index for Predicting Postoperative Moderate-to-Severe Pain: A Systematic Review and Meta-Analysis

**DOI:** 10.1101/2025.11.04.25339549

**Authors:** Wei Liu, Ying Li, Rong-Guo Yu, Han Chen, Qin Lin, Xiang-Feng Wang

## Abstract

**BACKGROUND:** Conventional vital signs lack the specificity for intraoperative nociception. The Surgical Pleth Index (SPI), calculated from photoplethysmographic waveforms, provides a quantitative measure of nociceptive status ranging from 0 to 100. Elevated SPI values correspond to increased nociceptive intensity. While some evidence suggests that SPI may help predict pain, its accuracy in forecasting postoperative pain requires further validation.

**AIM:** This study aimed to assess the capacity of the Surgical Pleth Index (SPI) to predict moderate to severe pain following surgery.

**METHODS:** We conducted a systematic literature search across three databases to identify studies investigating SPI’s predictive value for postoperative pain. A random-effects model was applied to pool summary estimates of sensitivity, specificity, and the area under the summary receiver operating characteristic curve (SROC-AUC).

**RESULTS:** Analysis included ten studies encompassing 1,042 patients. Pooled sensitivity and specificity were 0.74 (95% CI: 0.67–0.80) and 0.65 (95% CI: 0.55–0.74), respectively. The SROC-AUC reached 0.76, suggesting a moderate level of predictive accuracy. Significant heterogeneity was observed and not explained by differences in SPI cutoff values.

**CONCLUSION:** The SPI demonstrates moderate accuracy in forecasting moderate-to-severe postoperative pain and may serve as a useful adjunct to conventional clinical assessment.

**What is known?:** The Surgical Pleth Index has been suggested as a reliable monitor for nociceptive states.

**What new information does this article contribute?:** The Surgical Pleth Index (SPI) demonstrated moderate accuracy in predicting moderate-to-severe postoperative pain. Current evidence supports its role as a validated supplementary instrument to guide analgesic administration during surgery.

**Core Tip:**This meta-analysis confirms that the Surgical Pleth Index (SPI) provides moderate predictive accuracy for moderate-to-severe postoperative pain and, as such, has a complementary role in guiding intraoperative analgesia, provided its outputs are interpreted within the context of a comprehensive clinical assessment.

## INTRODUCTION

The neurophysiological processes that mediate pain perception are complex and remain poorly understood. Noxious stimuli typically evoke a sympathetic response that induces peripheral vasoconstriction, a physiological change that is directly correlated with the degree of sympathetic activation.

The Surgical Pleth Index (SPI) was calculated from the beat-to-beat interval and the amplitude of the photoplethysmographic pulse wave. It is a dimensionless parameter expressed on a calibrated scale, ranging from 0 to 100. Higher index values reflect an increased nociceptive intensity.^[1]^.

Prior investigations by Ledowski et al. ^[2–11]^ indicated the potential efficacy of the Surgical Pleth Index (SPI) in forecasting postoperative pain. The present meta-analysis consolidates this body of evidence to precisely quantify its diagnostic performance, thereby furnishing robust statistical evidence to validate its clinical utility in predicting moderate-to-severe postoperative pain.

## MATERIALS AND METHODS

### Protocol registration

The protocol for this meta-analysis was prospectively registered in PROSPERO (CRD42021250756).

### Review Question

This study aimed to assess the overall diagnostic performance of the Surgical Pleth Index (SPI) in predicting moderate-to-severe postoperative pain.

### Search Strategy and Terms

A systematic search was performed across PubMed, Cochrane Library, Embase, and China National Knowledge Infrastructure for records from inception to August 31, 2025. The search strategy, guided by the PICOS framework, encompasses a combination of keywords, Medical Subject Headings (MeSH), and free-text terms pertaining to general anesthesia, plethysmography, Surgical Pleth Index, Surgical Stress Index, hemodynamics, intraoperative monitoring, and acute pain. Unpublished and ongoing trials were identified using the WHO International Clinical Trials Registry Platform and ClinicalTrials.gov. The retrieved records were managed and screened using NoteExpress version 5.3.

### Inclusion and Exclusion Criteria

Studies that investigated the association between SPI monitoring and postoperative pain were included. The exclusion criteria were as follows: (1) reviews, commentaries, case reports, and animal studies; (2) articles from which a 2×2 contingency table could not be extracted; and (3) studies that presented only graphical data without accessible raw data.

### Selection Process

The initial screening of records was conducted independently by two investigators who removed duplicates and assessed titles and abstracts to determine eligibility for full-text retrieval.. Studies were included if a 2 × 2 table was directly available or calculable for the RevMan. Disagreements were resolved by discussion with a third reviewer.

### Data Extraction

The following data were extracted from each study: first author, publication year, journal name, country, ASA of Anesthesiologists physical status, patient age, reference standard, threshold value, sample size, and diagnostic metrics (including true positives, false positives, false negatives, true negatives, negative predictive values, positive predictive values, diagnostic odds ratios, sensitivity, and specificity). The corresponding authors were contacted for original data if the studies presented only graphical results.

### Ethical Considerations and Literature Search Overview

As this study constituted a secondary analysis of previously published literature without any direct involvement of human subjects, ethical approval from the institutional review board was not required. The search and methodology were performed according to Cochrane review standards.

### Statistical Analysis

Data for the 2×2 tables were derived using RevMan 5.3. A meta-analysis of dichotomous data was conducted using Stata 13.0. Heterogeneity was assessed using Cochran’s *Q* test and *I*² statistic. A fixed-effects model was employed when statistical heterogeneity was low (*p* ≥ 0.1 and *I*² ≤ 50%). Conversely, a random-effects model was adopted in the presence of significant heterogeneity. Derived summary estimates of sensitivity and specificity were computed using a random-effects approach. The overall diagnostic performance was evaluated by generating a summary receiver operating characteristic (SROC) curve, with the corresponding area under the curve (AUC) computed to quantify accuracy.

### Risk of Bias

The methodological quality of the included studies was independently appraised by two investigators using the QUADAS-2 tool in RevMan 5.3. Any discrepancies in the assessment were adjudicated through consultation with a senior researcher. Each study was categorized across four QUADAS-2 domains: Patient Selection, Index Test, Reference Standard, and Flow & Timing, as having a low, high, or unclear risk of bias.

### Meta-Regression

Considerable heterogeneity was anticipated, attributable to the expected variations across patient cohorts, surgical procedures, and pain assessment methodologies. A pre-specified meta-regression was conducted to investigate the potential sources of this heterogeneity, with a primary analytical focus on the SPI cutoff value. Additional covariates documented for this analysis included patient age, ASA of Anesthesiologists physical status, and the reference standard employed. The limited number of included studies (n=10) precluded meaningful multivariate meta-regression or subgroup analysis.

### Data Availability Statement

The datasets generated and analyzed during this meta-analysis can be obtained from the corresponding author upon legitimate request.

## RESULTS

### Search Process

A database search yielded 10,004 records. Following deduplication and screening of titles and abstracts, 72 publications advanced to the full-text assessment phase. Of these, ten articles fulfilled all the inclusion criteria and were included in the final meta-analysis (Figure 1).

**Figure 1.**
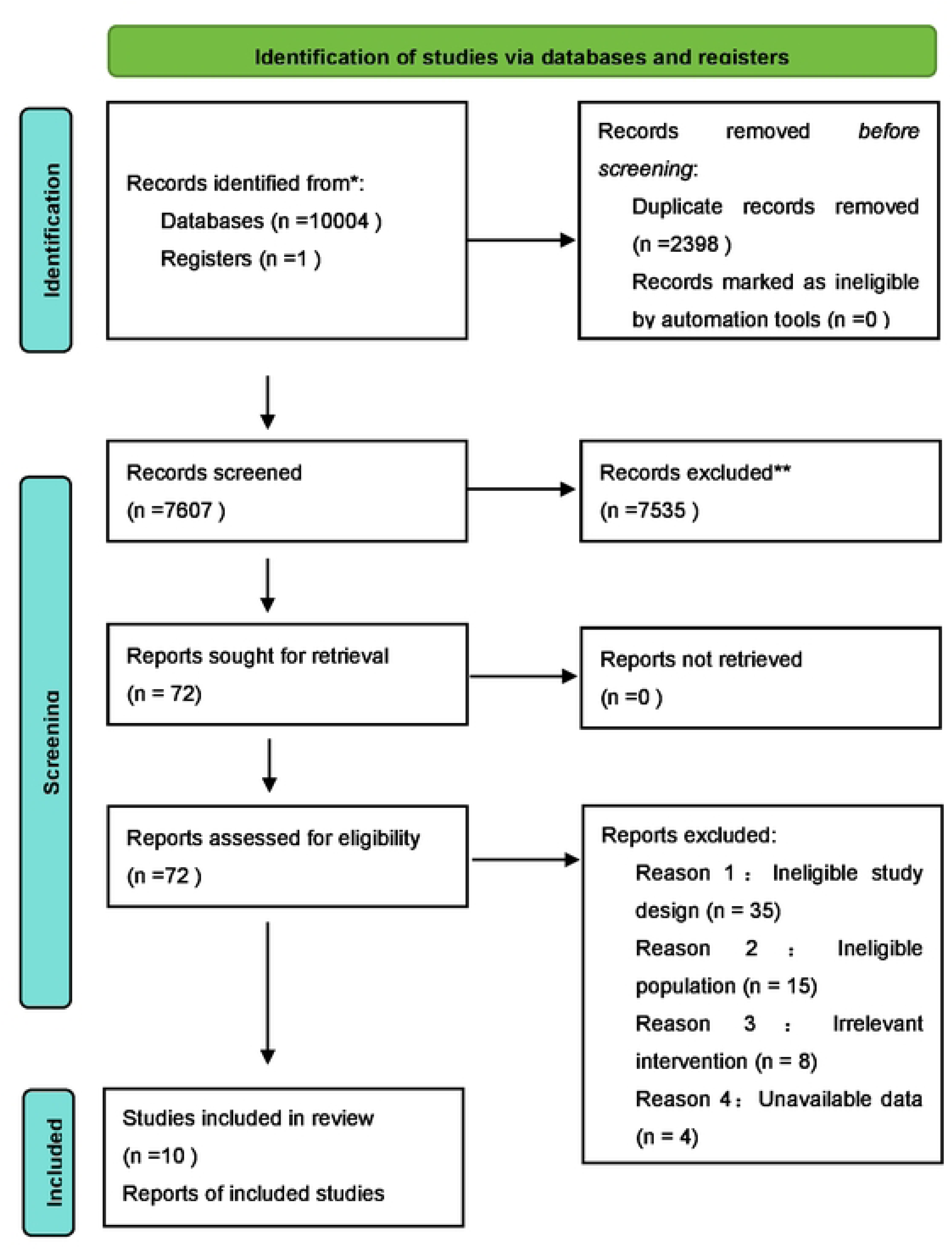
Flow chart of meta-analysis. 10004 records were identified by database searching. Following deduplication and screening of titles and abstracts, 72 articles advanced 10 full-text review. Of these, 62 were subsequently excluded for the following reasons: non-target population, unextractable data, outcome mismatch, or ineligible publication type. The remaining 10 studies satisfied all inclusion criteria and were incorporated into the meta-analysis.

Sixty-two articles were excluded due to non-target population, unextractable data (contact with authors for original data was unsuccessful for graphically presented studies), mismatched outcome measures, or ineligible publication type (e.g., review/meta-analysis).

### Study Characteristics

The final analysis included 10 studies (published between 2015 and 2024), encompassing a total of 1,042 patients (Table 1). The study sites were geographically diverse, spanning Australia, Germany, France, South Korea, China, and Mexico. Nine studies used the Numeric Rating Scale (NRS) as the reference standard, eight defined moderate-to-severe pain as NRS > 3, one as NRS > 5, and one used FLACC > 3 and FPS-R > 4. Considerable variation in SPI thresholds was observed, which likely contributed to high heterogeneity. Diagnostic data are presented in Table 2.

**Table 1.** Characteristics of included studies.

|  | Journal | Country/region | ASA | Age (yr) | Reference standard | Sample size |
| --- | --- | --- | --- | --- | --- | --- |
| 1.B.M. Choi<br>2018 | Anaesthesia | South Korea | I - II | 54.4 ± 12.9 | NRS>3 | 89 |
| 2. Dra. Mariel Alejandra<br>2024 | Revista Mexicana de Anestesiología | Mexico | I - II | 40.7 ± 15.5 | NRS>3 | 60 |
| 3.Chuan Jia 2021 | Master's Thesis, Hebei Medical University | China | I - II | 57 ± 9 | NRS>3 | 64 |
| 4.Ruijing Wang<br>2020 | Journal of Clinical Monitoring and Computing | China | I - II | 58.3 ± 13.6 | NRS>3 | 76 |
| 5.Carsten Thee<br>2015 | European Journal of Anaesthesiology | Germany | I - II | 57 ± 16 | NRS>3 | 100 |
| 6.T.Ledowski<br>2017 | British Journal of Anaesthesia | Australia | I - II | 2-16 | FLACC>3<br>FPS-R>4<br>NRS>3 | 93 |
| 7.T.Ledowski<br>2016 | British Journal of Anaesthesia | Australia | -- | 43 ± 15 | NRS>31 | 65 |
| 8.T.Bapteste<br>2018 | Anaesthesia, Critical Care & Pain Medicine | France | -- | -- | VAS>3 | 250 |
| 9.T.Ledowski<br>2019 | British Journal of Anaesthesia | Australia | -- | 18-95 | NRS>3 | 196 |
| 10.MiHye Park<br>2020 | Journal of Clinical Monitoring and Computing | South Korea | I -II | 68 ± 2 | NRS>5 | 49 |

**Table 2.**
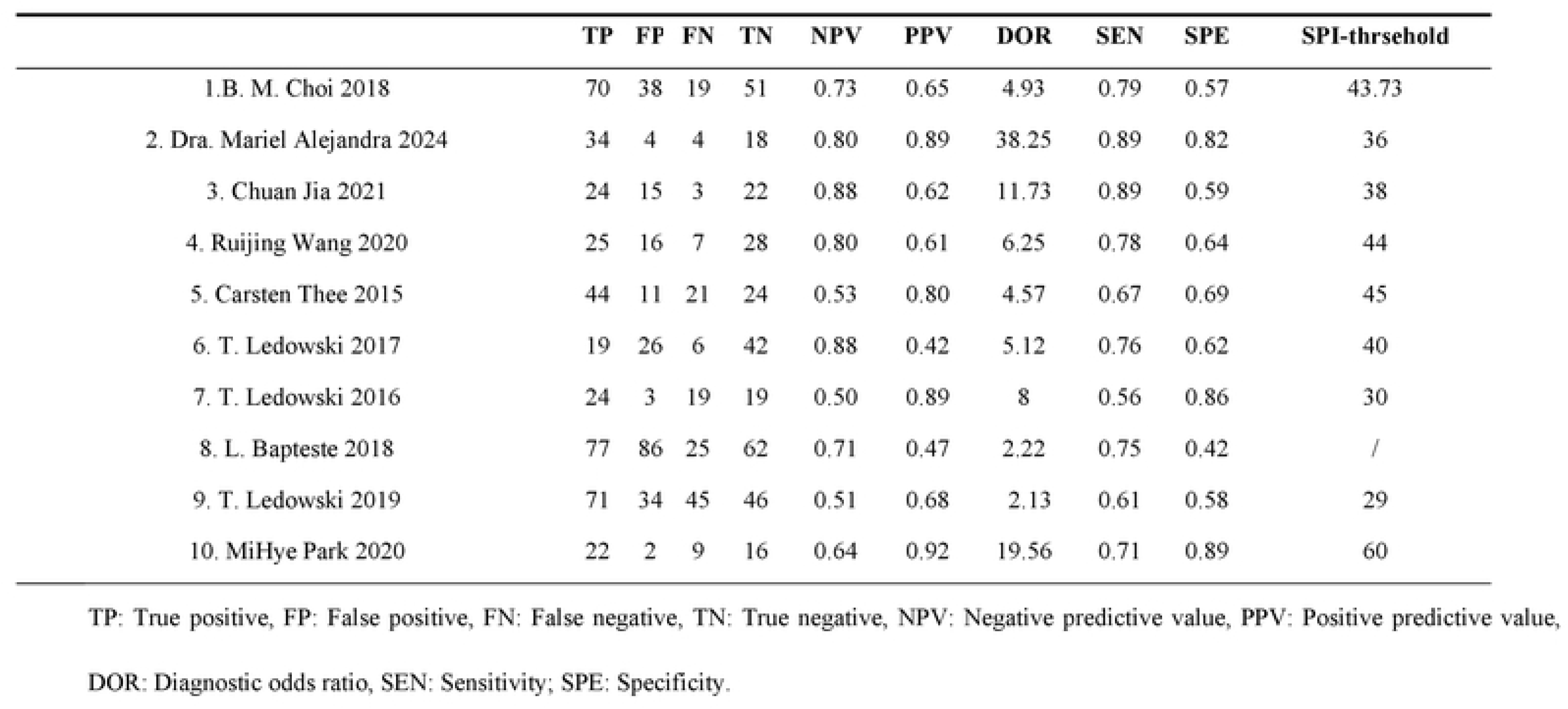
Diagnostic parameters of the included studies.

|  | **TP** | **FP** | **FN** | **TN** | **NPV** | **PPV** | **DOR** | **SEN** | **SPE** | **SPI-thrsehold** |
| --- | --- | --- | --- | --- | --- | --- | --- | --- | --- | --- |
| 1.B. M. Choi 2018 | 70 | 38 | 19 | 51 | 0.73 | 0.65 | 4.93 | 0.79 | 0.57 | 43.73 |
| 2. Dra. Mariel Alejandra 2024 | 34 | 4 | 4 | 18 | 0.80 | 0.89 | 38.25 | 0.89 | 0.82 | 36 |
| 3. Chuan Jia 2021 | 24 | 15 | 3 | 22 | 0.88 | 0.62 | 11.73 | 0.89 | 0.59 | 38 |
| 4. Ruijing Wang 2020 | 25 | 16 | 7 | 28 | 0.80 | 0.61 | 6.25 | 0.78 | 0.64 | 44 |
| 5. Carsten Thee 2015 | 44 | 11 | 21 | 24 | 0.53 | 0.80 | 4.57 | 0.67 | 0.69 | 45 |
| 6. T. Ledowski 2017 | 19 | 26 | 6 | 42 | 0.88 | 0.42 | 5.12 | 0.76 | 0.62 | 40 |
| 7. T. Ledowski 2016 | 24 | 3 | 19 | 19 | 0.50 | 0.89 | 8 | 0.56 | 0.86 | 30 |
| 8. L. Bapteste 2018 | 77 | 86 | 25 | 62 | 0.71 | 0.47 | 2.22 | 0.75 | 0.42 | / |
| 9. T. Ledowski 2019 | 71 | 34 | 45 | 46 | 0.51 | 0.68 | 2.13 | 0.61 | 0.58 | 29 |
| 10. MiHye Park 2020 | 22 | 2 | 9 | 16 | 0.64 | 0.92 | 19.56 | 0.71 | 0.89 | 60 |

TP: True positive, FP: False positive, FN: False negative, TN: True negative, NPV: Negative predictive value, PPV: Positive predictive value, DOR: Diagnostic odds ratio, SEN: Sensitivity; SPE: Specificity.

### Patient Characteristics

The population included adults (mean age, ∼68 years) and pediatric patients (2–16 years) classified as ASA I–III. Considerable heterogeneity was noted in the predictive accuracy among the included studies, with sensitivity estimates ranging from 56% to 89%, and specificity values ranging from 42% to 89%. For instance, Ledowski et al. (2016) documented a lower sensitivity of 56% coupled with a higher specificity of 86%. In contrast, a more recent investigation by Alejandra et al. (2024) reported a high sensitivity of 89% and specificity of 82%. The observed heterogeneity likely originates from inter-study differences in surgical procedures, applied SPI thresholds, pain assessment criteria, and limited sample sizes of certain cohorts. The detailed baseline characteristics and diagnostic parameters are summarized in Tables 1 and 2, respectively.

### Risk of Bias

Figure 2 presents the methodological quality assessment of the ten included studies, as evaluated using the QUADAS-2 tool. All investigations recruited patients experiencing postoperative pain and employed SPI monitoring and a validated reference standard for diagnostic confirmation.

**Figure 2.**
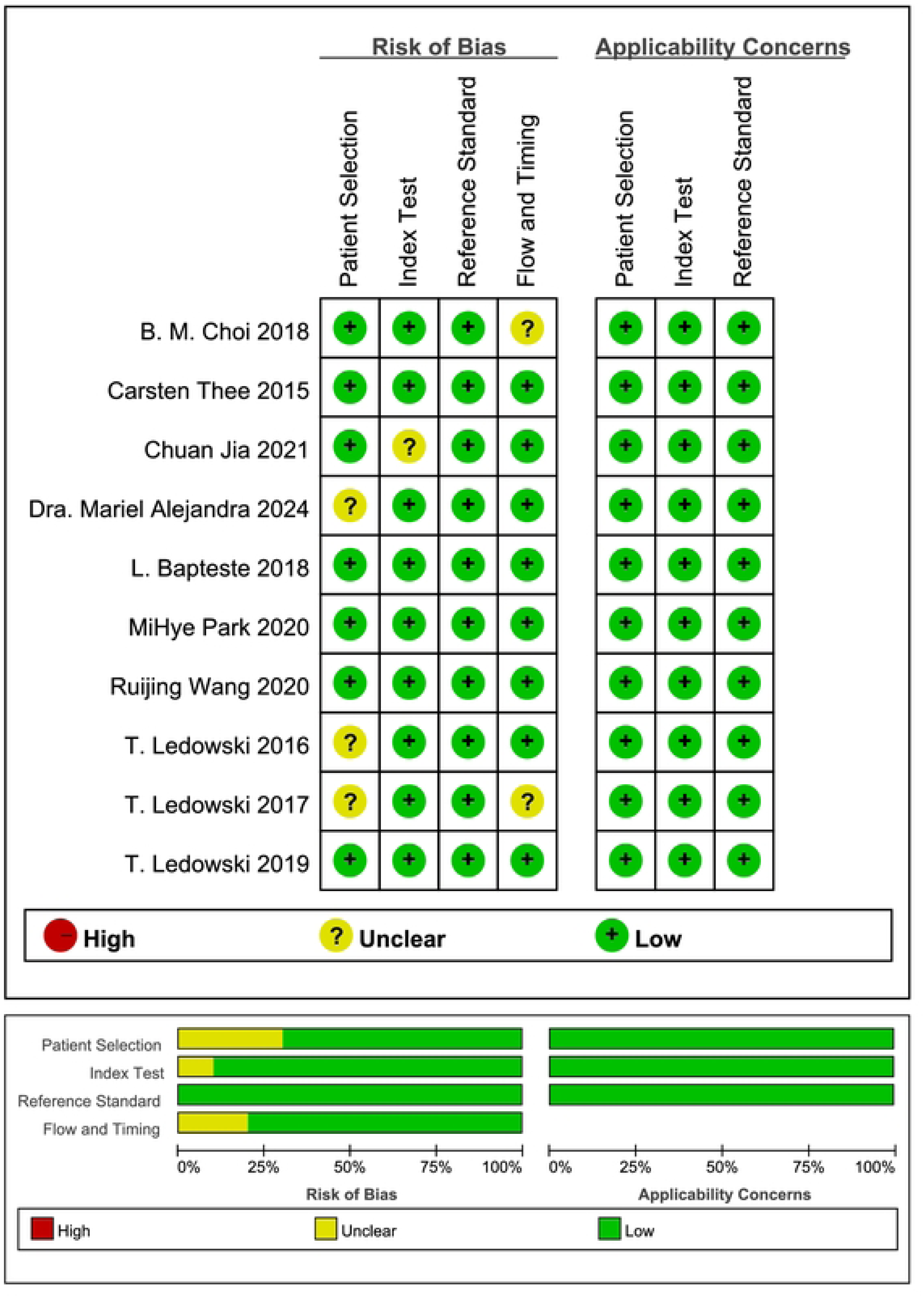
Risk of bias assessment of the 10 included studies. The methodological quality of the 10 included studies, evaluated using the QUADAS-2 tool, demonstrated variable risk of bias across four key domains: patient selection, index test, reference standard. and flow/timing. The assessments were categorized as high, low, or unclear.

### Main Results of Meta-Analysis

A random-effects model was used. Significant heterogeneity was observed for sensitivity (*Q*=29.10, *I*²=69.07%, df=9, *p*<0.001; Figure 3) and specificity (*Q*=40.98, *I*²=78.04%, df=9, *p*<0.001). Figure 4 displays the bivariate boxplot of the logit-transformed sensitivity and specificity, revealing considerable dispersion among the included studies. The surrounding gray 95% confidence region reflects moderate diagnostic consistency, despite this heterogeneity. Summary receiver operating characteristic (SROC) analysis yielded an area under the curve (AUC) of 0.76 (95% CI: 0.09–0.99; Figure 5). In the Fagan nomogram, assuming a pre-test probability of 20%, a positive SPI result increased the post-test probability to 35% (positive likelihood ratio [LR+] = 2), whereas a negative result reduced it to 9% (negative likelihood ratio [LR−] = 0.40), indicating clinically meaningful, though modest, probability shifts (Figure 6). The likelihood ratio scatter plot positioned the summary LR+ (2.0) and LR− (0.40) in the lower-left quadrant, supporting the utility of SPI primarily for ruling out, rather than confirming, moderate-to-severe postoperative pain (Figure 7). Meta-regression identified no statistically significant relationship between the SPI threshold and the negative predictive value (coefficient = 0.013, 95% CI: –0.060 to 0.087, *p* = 0.680). Considerable between-study heterogeneity remained (*I*² = 77.06%), with the SPI cutoff explaining only a negligible proportion of the variance (adjusted R² = –13.46%; Table 3).

**Figure 3.**
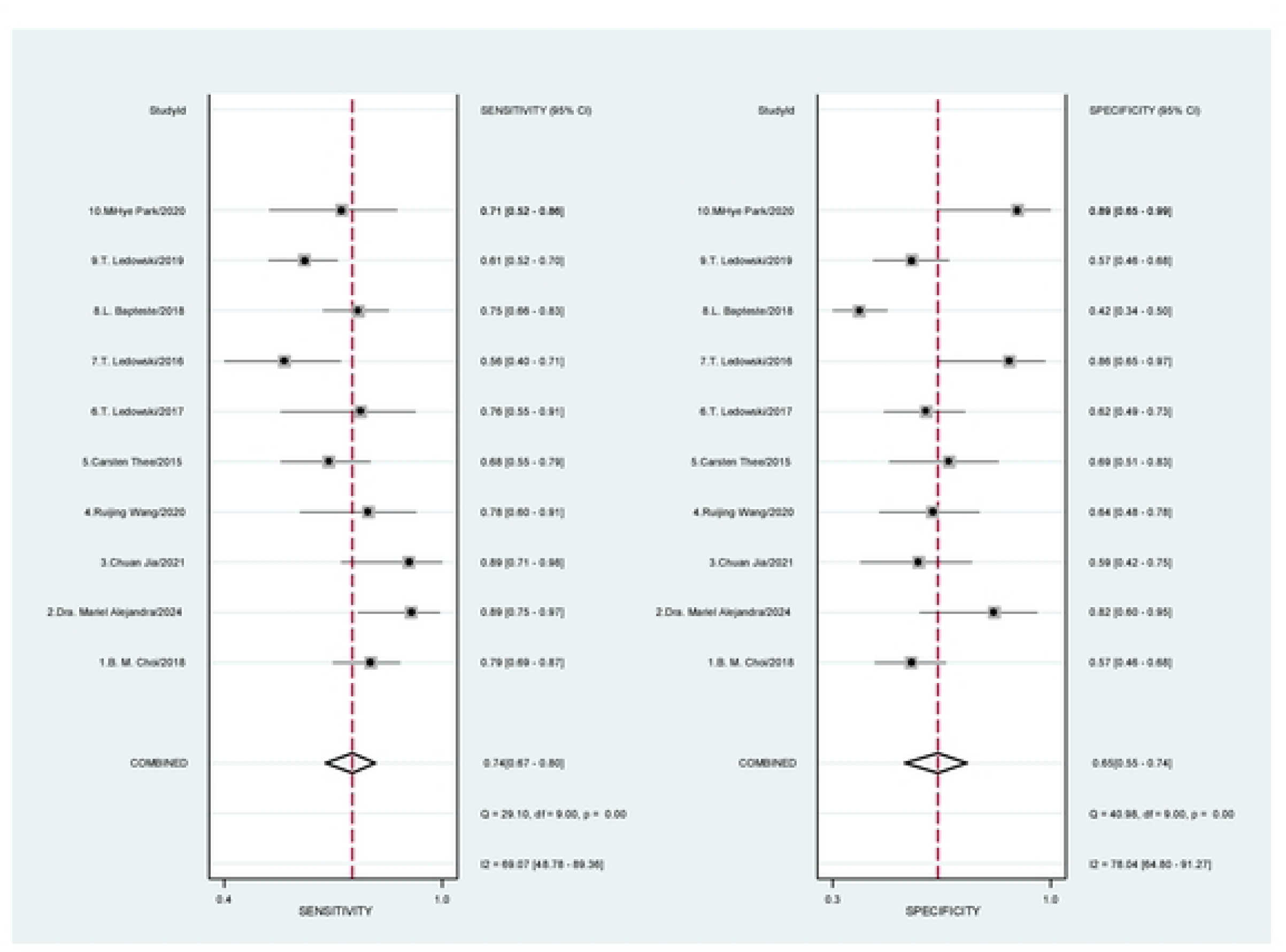
Sensitivity and specificity of the included studies. The left forest plot shows sensitivity in the 10 studies. Surgical Pleth Index (SPI) predicted postoperative pain. Each line refers to one of the studies. The diamond at the bottom shows the pooled result. The pooled sensitivity was 0.74. Significant heterogeneity among the studies was observed (I^2^ = 69.07%). The right forest plot shows specificity. The pooled specificity was 065. There was significant variation between studies (I^2^ = 78.04%).

**Figure 4.**
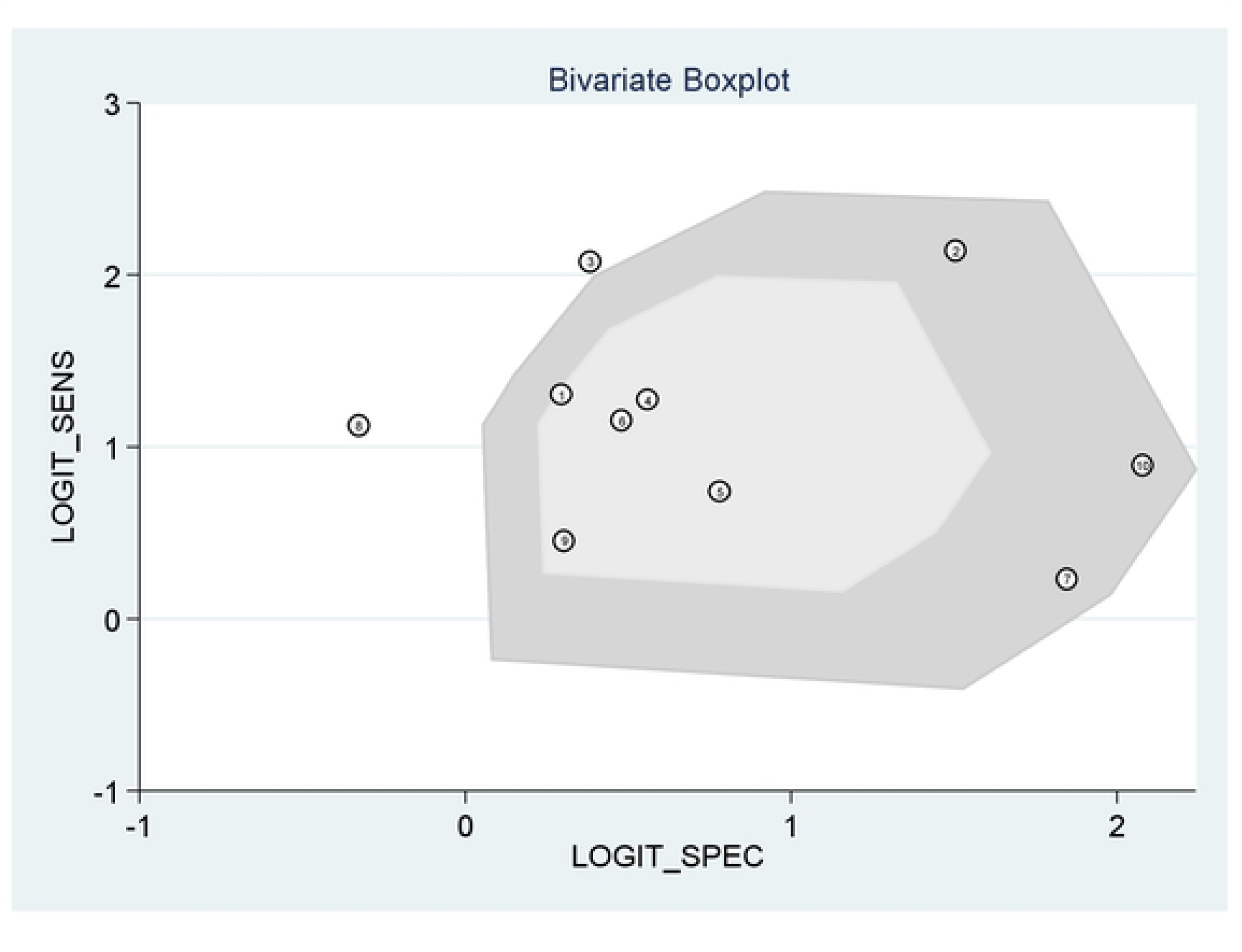
Bivariate boxplot of logit-sensitivity and logit-specificity for the Surgical Pleth Index (SPJ) in predicting postoperative pain. This was a bivariate boxplot that showed the spread of data. It used two measures: LOGIT_SENS and LOGIT_SPEC. These were adjusted sensitivity and specificity values. The data were taken from multiple SPI studies. Each circle represents a study. The gray area shows the 95% confidence range. This plot summarized the diagnostic performance of SPI and showed the spread of the data and where most values cluster.

**Figure 5.**
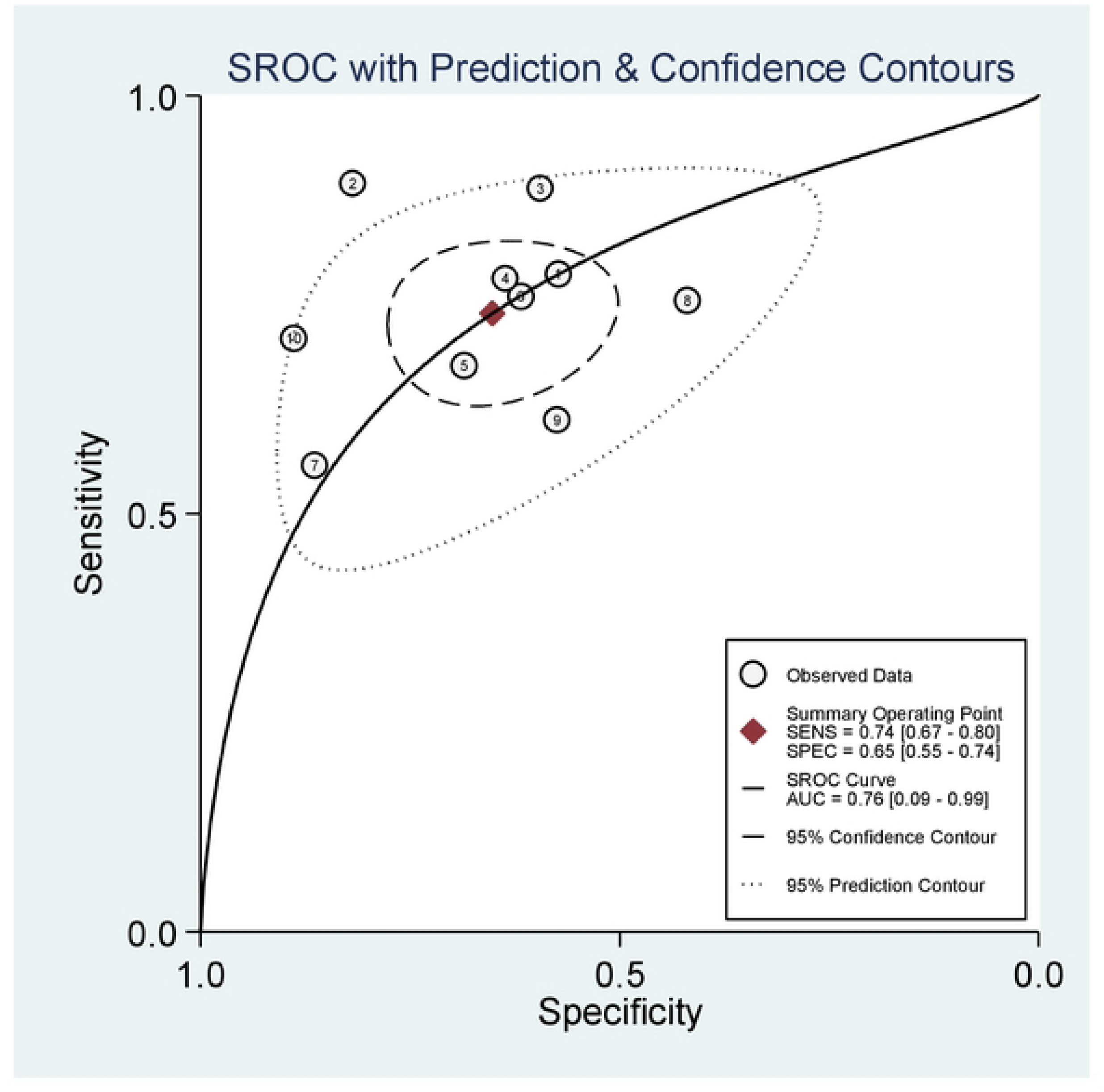
Summary receiver operating characteristic (SROC) curve with prediction and confidence contours. SPI predicted postoperative pain. Each circle represents a study. The bigger circles indicate larger sample sizes. The solid line was the SROC curve. Area under the curve (AUC) was 0.76, which indicates moderate accuracy. The red diamond shows the summary point. Dashed lines show the 95% confidence range. Dotted lines show the 95% prediction range.

**Figure 6.**
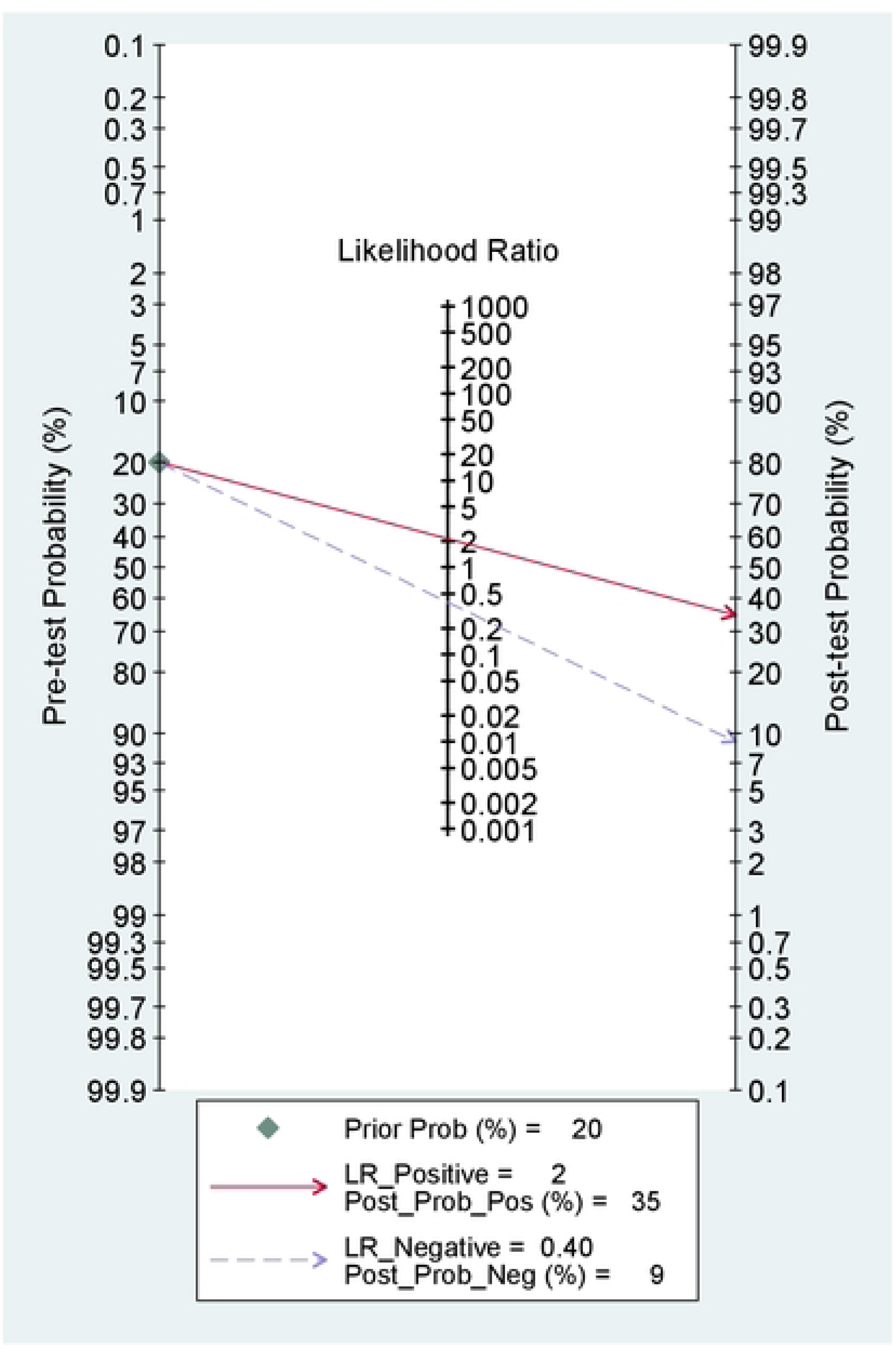
Fagan diagram illustrating the impact of a diagnostic test on post-test probability based on pre-test probability and likelihood ratios. The Fagan plot showed how pre-test probability and likelihood ratios affected post-test probability. The red solid line was the positive likelihood ratio (LR+), which was 2. A positive test doubled the disease probability. The blue dashed line was the negative likelihood ratio (LR−), which was 0.40. A negative test halved the disease probability. The diamond marker showed a prior probability of 20%. The probability became 35% after a positive test and 9% after a negative test.

**Figure 7.**
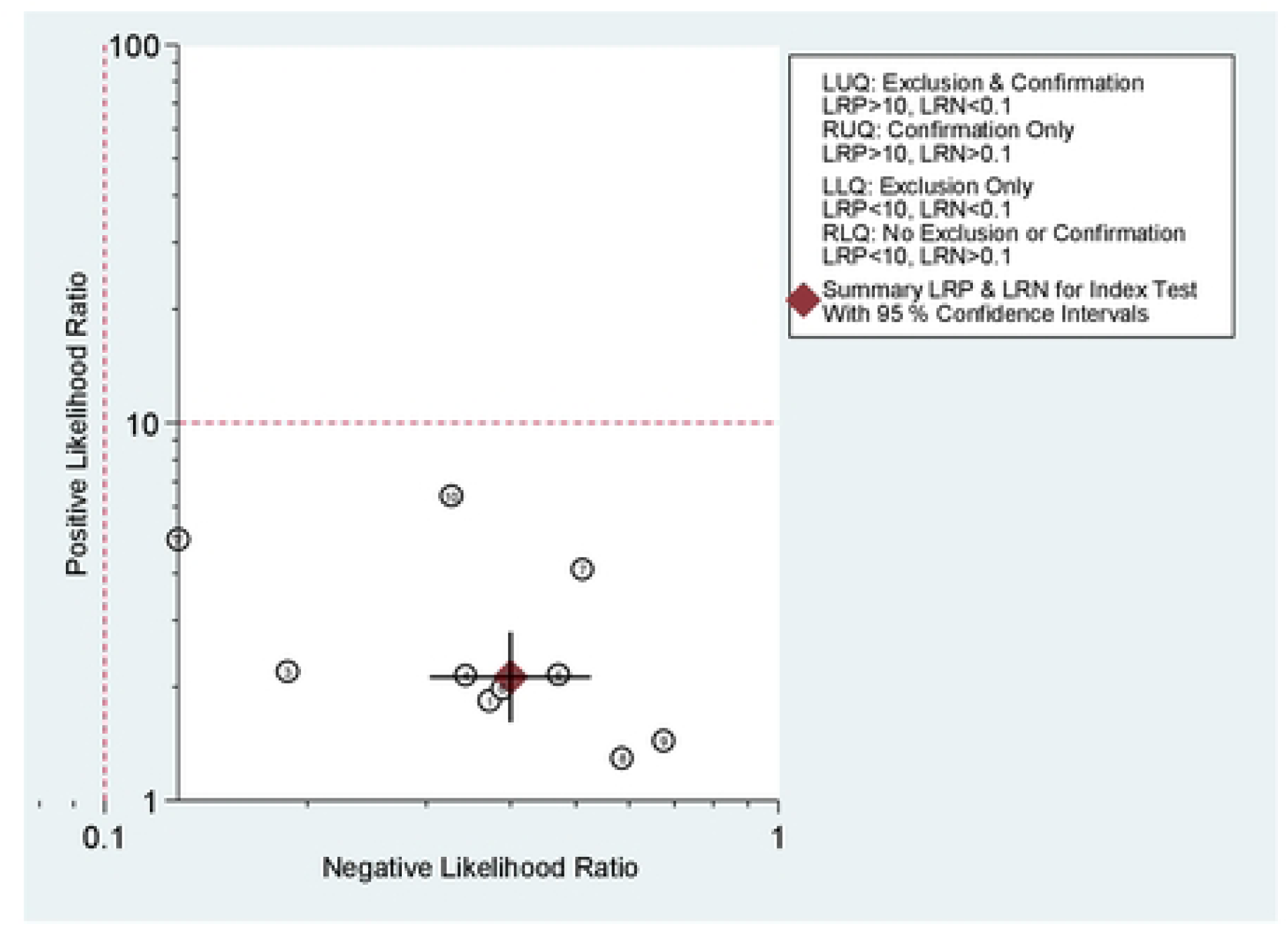
Likelihood ratio scatter plot for the Surgical Pleth Index (SPI) in predicting postoperative pain. The scatter plot showed how SPI predicted postoperative pain. It used positive and negative likelihood ratios (LR+ and LR−). One circle represents one study. The diamond shows the overall results with 95% confidence intervals. Red dashed lines show LR+ = 10 and LR− = 0.1. These lines divided the plot into four sections. Each section showed how useful the test was: Top-left (very good at ruling in and ruling out); top-right (only good at ruling in); bottom-left (only good at ruling out) and bottom-right (not good at ruling in or out). The overall result was in the bottom-left section. This showed that SPI was better at ruling out pain than ruling it in. The performance was moderate.

**Table 3.** Meta-Regression Analysis of SPI Threshold.

| **Variable** | **Coefficient** | **Standard Error** | **t-value** | **P-value** | **95% Confidence Interval** |
| --- | --- | --- | --- | --- | --- |
| SPI Threshold | 0.013 | 0.031 | 0.43 | 0.680 | -0.060 to 0.087 |
| Intercept | 0.321 | 1.290 | 0.25 | 0.811 | -2.730 to 3.371 |

A random-effects meta-regression model was used. The Knap–Hartung adjustment was applied to the confidence intervals.

## DISCUSSION

Postoperative pain is a common clinical challenge and substantial source of patient morbidity^[12]^. A key objective of perioperative care is to optimize the administration of analgesics during surgery, thereby preventing the onset and reducing the intensity of postoperative pain.. Prior research, particularly studies by Ledowski et al. ^[2–11]^, has suggested the potential of the Surgical Pleth Index (SPI) as a predictive tool. Therefore, this meta-analysis was designed to synthesize reliable evidence for the diagnostic accuracy of the Surgical Pleth Index (SPI). Determining its clinical value could facilitate enhanced pain management, promote faster recovery, reduce complication rates, and improve overall patient outcomes.

Conventional assessment of nociception is often dependent on physiological parameters, such as heart rate and blood pressure, which are indirect surrogate markers and lack specificity. Heart rate variability (HRV) captures oscillations in the beat-to-beat interval; however, it does not directly correlate with pain intensity and does not permit quantitative comparisons of pain levels between individuals^[13]^. Although photoplethysmography (PPG) detects peripheral blood volume fluctuations linked to nociception, its clinical application generally depends on sophisticated signal-processing techniques^[14]^. A computational model that integrates PPG with HRV data, further calibrated to minimize inter-individual variability, has demonstrated improved specificity for pain assessment. The resulting SPI formula weighs PPG more heavily: SPI = 100 – (0.33 × HRV + 0.67 × PPG).

The causal link between the Surgical Pleth Index (SPI) and postoperative pain is supported by its underlying physiology. SPI provides a real-time reflection of sympathetic tone, which is elevated by nociceptive stimuli under conditions of inadequate analgesia^[15–17]^. This heightened sympathetic activity signifies the pain-induced sensitization of the central nervous system. A sensitized state can extend into the postoperative period, whereby elevated intraoperative SPI values, indicative of a hypersensitive nervous system, predict an increased risk of moderate-to-severe pain^[18–20]^. The pooled AUC of 0.76 derived from our meta-analysis confirms the good discriminatory power of SPI for this outcome, thereby validating its predictive utility. In summary, SPI captures a neurophysiological state of pain sensitization driven by sustained nociceptive input, and an AUC exceeding 0.7 substantiates its value as a predictive tool.

Considerable statistical heterogeneity was observed across the included studies (*I*² > 75%). Despite the initial assumptions that varying SPI thresholds might be the primary cause, meta-regression analysis did not identify threshold variation as a statistically significant contributor. This implies that establishing a universal SPI cutoff may be unnecessary and that the observed heterogeneity likely arises from legitimate clinical and methodological diversity. The key sources of variation included divergent patient demographics (encompassing both adult and pediatric populations, wherein age influences autonomic reactivity and pain perception), dissimilar surgical procedures (each eliciting distinct nociceptive profiles), inconsistent application of pain assessment tools (NRS, VAS, and FLACC), differing definitions of moderate-to-severe pain (NRS > 3 vs. NRS > 5), heterogeneous ASA physical status distributions, and varying sample sizes (with smaller studies producing less precise estimates). A random effects model was used to derive a conservative summary estimate. The SROC curve, with an AUC of 0.76, corroborates the clinical value of the SPI for postoperative pain monitoring. Future investigations should prioritize the development of standardized assessment protocols and validate SPI performance within more homogeneous patient cohorts to improve reliability and generalizability.

This systematic review had several limitations. The significant heterogeneity observed likely stems from differences in SPI threshold selection, diversity of surgical interventions, and variations in pain assessment methodologies.. Furthermore, the limited number of eligible studies precluded a more thorough investigation through meta-regression or subgroup analyses, as such analyses would have been underpowered. No formal assessment of publication bias was conducted because robust and validated methods specifically tailored for diagnostic test accuracy meta-analyses are currently lacking.

While prior investigations have established the prognostic utility of the Surgical Pleth Index (SPI) measured at the end of surgery for predicting postoperative pain ^[19]^, the present study specifically examined intraoperative SPI trends and correlated these fluctuations with postsurgical pain outcomes. This methodological approach offers a more refined assessment of nociception-antinociception balance, thereby enabling the identification of dynamic pain patterns throughout the surgical procedure and potentially facilitating more personalized analgesic management. Consequently, this study provides an innovative evidence base for the field, although subsequent studies are required to standardize and validate SPI-guided analgesic protocols.

## CONCLUSION

The Surgical Pleth Index is a useful tool for predicting postoperative pain with moderate accuracy.

## Author contributions

**Study design and conception:** Wei Liu, Rong-Guo Yu, Han Chen, Xiang-Feng Wang.

**Implementation and data collection:** Wei Liu, Qin Lin, Xiang-Feng Wang.

**Statistical analysis:** Wei Liu, Ying Li, Xiang-Feng Wang.

**Manuscript – initial drafting:** Wei Liu, Ying Li, Xiang-Feng Wang.

**Manuscript – critical revision and final approval:** Wei Liu, Rong-Guo Yu, Han Chen, Xiang-Feng Wang.

**Overall supervision and quality control:** Wei Liu, Xiang-Feng Wang.

All authors reviewed and approved the final manuscript.

## Author contributions

The author contributions section from the original manuscript is retained verbatim. Conflict of interest: The authors declare no conflicts of interest.

## Data availability statement

Data are available upon reasonable request. Ethical approval: Not applicable.

## Informed consent

Not applicable.

## Acknowledgment

The authors acknowledge the use of DeepSeek (https://www.deepseek.com/) to assist in the polishing of language and grammar in parts of this manuscript. It was also used as a tool for brainstorming during the initial structuring of the discussion. All output generated by the AI was critically reviewed and modified by the authors. The authors assume full responsibility for the accuracy, integrity, and ethical compliance of the final content.

